# Antihypertensive medications and dementia in older adults with hypertension

**DOI:** 10.1101/2024.08.28.24312754

**Authors:** Suzanne G. Orchard, Zhen Zhou, Michelle Fravel, Joanne Ryan, Robyn L. Woods, Rory Wolfe, Raj C. Shah, Anne Murray, Ajay Sood, Christopher M. Reid, Mark R. Nelson, Lawrie Bellin, Kevan R Polkinghorne, Nigel Stocks, Michael E. Ernst

**Affiliations:** School of Public Health and Preventive Medicine, Monash University, Melbourne, VIC, Australia; Menzies Institute for Medical Research, University of Tasmania, Hobart, TAS, Australia; Department of Family Medicine, Carver College of Medicine, The University of Iowa, Iowa City, Iowa, USA; Department of Family and Preventive Medicine and Rush Alzheimer’s Disease Center, Rush University Medical Center, Chicago, USA; Berman Center for Outcomes and Clinical Research, Hennepin Healthcare Research Institute, and Division of Geriatrics, Department of Medicine Hennepin HealthCare, Minneapolis; University of Minnesota, Minneapolis, USA; Department of Neurology and the Rush Alzheimer’s Disease Center, Rush University Medical Center, Chicago, Illinois, USA; School of Public Health, Curtin University, Perth, WA, Australia; Medical School Royal Perth Hospital, University of Western Australia, Perth, WA, Australia; Department of Nephrology, Monash Medical Centre, Monash Health, Melbourne, VIC, Australia; Department of Medicine, Monash University, Melbourne, VIC, Australia; Discipline of General Practice, Adelaide Medical School, University of Adelaide, SA, Australia; Department of Pharmacy Practice and Science, College of Pharmacy, The University of Iowa, Iowa City, Iowa, USA

**Keywords:** Antihypertensive medication, Angiotensin II receptor blocker, Angiotensin-converting enzyme inhibitor, dementia, cognition, aged, older adults

## Abstract

**Background:** Studies on middle-aged or individuals with cognitive or cardiovascular impairments, have established that intensive blood pressure (BP) control reduces cognitive decline risk. However, uncertainty exists on differential effects between antihypertensive medications (AHM) classes on this risk, independent of BP-lowering efficacy, particularly in community-dwelling hypertensive older adults.

**Methods:** A post-hoc analysis of the ASPREE study, a randomized trial of low-dose aspirin in adults aged 70+ years (65+ if US minorities) without baseline dementia, and followed for two years post-trial. Cox proportional-hazards regression models were used to estimate associations between baseline and time-varying AHM exposure and incident dementia (an adjudicated primary trial endpoint), in participants with baseline hypertension. Subgroup analyses included prespecified factors, APO ε4 carrier status and monotherapy AHM use.

**Results:** Most hypertensive participants (9,843/13,916; 70.7%) used AHMs. Overall, ‘any’ AHM use was not associated with lower incident dementia risk, compared with untreated participants (HR 0.84, 95%CI 0.70-1.02, p=0.08), but risk was decreased when angiotensin receptor blockers (ARBs) were included (HR 0.73, 95%CI 0.59-0.92, p=0.007). ARBs and β-blockers decreased dementia risk, whereas angiotensin-converting enzyme inhibitors (ACEIs) and diuretics increased risk. There was no association with RAS modulating or blood-brain-barrier crossing AHMs on dementia risk.

**Conclusions:** Overall, AHM exposure in hypertensive older adults was not associated with decreased dementia risk, however, specific AHM classes were with risk direction determined by class; ARBs and β-blockers were superior to ACEIs and other classes in decreasing risk. Our findings emphasize the importance of considering effects beyond BP-lowering efficacy when choosing AHM in older adults.

## INTRODUCTION

Dementia affects an estimated 55 million individuals globally, and expected to reach 70 million and 139 million by 2030 and 2050, respectively.^1^ The risk and prevalence of dementia increases with age, without an apparent ceiling.^2^ Hypertension, one of the most prevalent and modifiable risk factors for dementia,^3^ also increases in prevalence with age,^4^ leading to older adults accounting for the bulk of hypertension-related morbidity and mortality.^5^ Hypertension can directly impact cognition through increased risk for stroke and subsequent post-stroke cognitive impairment, ^6^ but also indirectly by hypertension-related changes in the cerebral vasculature, impairing cerebral perfusion and inducing inflammation and tissue damage.^7^ The treatment of hypertension in older adults is therefore a public health priority.

Despite existing comprehensive reviews on hypertension’s pathophysiology and links to dementia,^8–10^ evidence on the relationship between hypertension, treatment strategies and dementia in the very old (>80 years) remains limited^8^ and inconsistent.^10,11^ These inconsistencies are partly driven by the study population (e.g., cognitively intact, mildly impaired or with dementia or CVD), but also by the varying effects of individual antihypertensive medications (AHM) classes on dementia, independent of their blood-lowering effect.^10,12–15^ For example, prior studies found that renin-angiotensin system (RAS) inhibitors, (angiotensin II receptor type 1 (AT1) blockers (ARBs) and angiotensin-converting-enzyme inhibitors (ACEIs)), had the potential to reduce dementia risk or progression from mild cognitive impairment to dementia,^13,15^ whilst many have reported no clear association.^16,17^ It remains unclear whether the association its directionality, with the development and progression of dementia differs between AHM class, especially ACEIs and ARBs,^18–22^ in older cognitively intact adults.

Using the comprehensive cognitive and medication use data collected in the ASPirin in Reducing Events in the Elderly (ASPREE) trial^23,24^ and extended 2 years post-trial period, ASPREE-XT,^25^ we conducted a post-hoc analysis in community-dwelling, cognitively intact older adults who had hypertension at trial entry, to determine: (1) the associations of baseline and time-varying AHM use with long-term incident all-cause dementia; (2) the role of AHM class, including RAS modulation or blood-brain-barrier penetrance, in mediating AHM-related dementia risk; and (3) the extent that mono- and/or combination-AHM use, or APOE ε4 carrier status, were associated with changes in the risk profile.

## METHODS

### Data Availability and Study Population

The authors declare that all supporting data are available within the article, and its online supplementary files. Researcher access to the ASPREE study longitudinal dataset is via application through the ASPREE data access management system (https://ams.aspree.org/application/home.aspx).

This is a post-hoc analysis of data from the ASPREE study. Details on study design, recruitment, and baseline population characteristics at clinical trial enrolment and entry into the observational extension (ASPREE-XT) have already been described.^23–25^ ASPREE was a prospective, randomized placebo-controlled trial comparing effects of daily low-dose aspirin (100mg) versus placebo, on disability-free survival. Briefly, 19,114 healthy, community-dwelling people aged ≥70 years (≥65 years for US minorities) were randomized in Australia (n=16,703, 87%) and the US (n=2,411, 13%) from March 2010 to December 2014. The trial concluded in June 2017 (median follow-up 4.7 years, IQR, 3.6-5.7) and involved annual in-person study visits between 2011 and 2017. Eligible participants were free from evidence of cardiovascular disease (CVD), independence-limiting physical disability, and expected to survive for at least 5 years. Individuals with persistent severe hypertension (defined as ≥180 and/or ≥105 mmHg), a self-report or physician diagnosis of dementia, or a Modified Mini-Mental State Examination (3MS)^26^ score of <78/100, were ineligible. Participants provided informed consent and local ethics committees approved the study.

Participants with known baseline hypertension (HTN; defined as systolic/diastolic blood pressure [SBP/DBP] of <180 -≥140 and/or <105 -≥90 mmHg and/or self-report of AHM use) were included in this analysis and followed through to the second study visit post-trial (observational phase), median [IQR] follow-up: 6.4 [5.3-7.6] years).

### Exposure to Antihypertensive Medications (AHMs)

Participants brought all currently used medications, a list or self-report of medications, to annual visits, with subsequent confirmation via primary care practice medical records. Participants were sorted by baseline AHM use (treated vs untreated) and by AHM class. The ‘untreated’ group comprises those with HTN who did not use any AHM at baseline. AHMs were classified according to their primary mode of action: ARBs; ACEI; diuretics; Calcium Channel Blockers (CCBs); and β-blockers (BBs). Other, less frequently used AHMs (n=549, ATC codes beginning with ‘C02’) were excluded from analyses by class, but included in combination therapy. AHMs were also categorized by whether they were RAS stimulating or inhibiting.^20^ Determination of blood-brain-barrier (BBB)-crossing potential was established following previous literature.^27^

### Ascertainment of dementia

All-cause incident dementia was defined according to the Diagnostic and Statistical Manual for Mental Disorders, 4^th^ edition (DSM-IV) criteria, ^28^ and details previously outlined.^24^ Briefly, suspected cases of dementia (3MS score ≤77/100 or drop of >10.15 points from the predicted 5-year age- and education-adjusted score, reported cognitive concerns in medical records, a clinician diagnosis of dementia, or prescription of cholinesterase inhibitors) were referred for further cognitive assessment and then adjudicated by an expert panel blinded to treatment assignment.

### Assessment of covariates

Baseline confounders were selected based on known associations with dementia and potential interaction effects with AHMs.^29^ Model 1 adjusted for age and sex, and Model 2 further adjusted for race/ethnicity (White, Black, Hispanic/Latino, Others [includes Australian aborigine/Torres Strait islander, native American, more than one race, native Hawaiian/Pacific Islander and non-Hispanics who did not state their ethnicity/race]), country (Australia, US), years of education (<12, ≥12 years), smoking status (never, former, current), alcohol consumption (never, former, current), mean total cholesterol, living alone (yes/no), polypharmacy (≥5 prescription medications), family history of dementia (self-report), chronic kidney disease (CKD; estimated glomerular filtration rate <60 ml/min/1.73m^2^ or urinary albumin-to-creatinine ratio ≥3mg/mmol), diabetes (self-report or fasting glucose ≥ 126mg/dL or glucose-lowering medication use), statin use, baseline composite cognitive z scores (see Appendix S1 for calculation), depression score (8+/30) measured by the Center for Epidemiologic Studies Depression 10-item Scale (CES-D 10),^30^ baseline SBP and DBP, and ASPREE randomized treatment (aspirin/placebo). Procedures for covariate assessments are described in the ASPREE protocol (https://aspree.org/). Results are discussed for the fully adjusted model (Model 2), unless otherwise indicated.

### Statistical analyses

Descriptive statistics (frequencies (%) and means (SDs)) were used to summarize AHM prevalence in the ASPREE-HTN population. Cox proportional-hazards regression models were used to estimate the association between baseline AHM exposure (by ‘any’ exposure [yes/no], by monotherapy and/or combination therapy, by class [ARBs, ACEI, diuretics, CCB, BBs] and by RAS-stimulating or -inhibiting capability and BBB penetrance) and incident dementia. Participants were followed until the occurrence of dementia, death, or end of follow-up, whichever occurs first. Proportional-hazards assumptions were checked using Schoenfeld residuals test and no violations were detected. We repeated all analyses for incident dementia by treating AHM use for each class as a time-varying variable. Sensitivity analyses were conducted in a sub-cohort with known APOE ε4 genotype carrier status (10,538 genotyped; with 2684 ε4 carriers), further adjusting Model 2 for APOE ε4 status (Model 3), which did not change the main findings. We also repeated the main analysis with a 2-year lag period excluding participants with follow-up time or incident dementia <2 years from the baseline to avoid the potential reverse causation. All p-values were two-sided and p<0.05 was considered as significant. Analyses were performed using Stata/SE (StataCorp, version 15.0).

## RESULTS

Participants without HTN (n=4,919) and those with missing values for baseline composite cognitive test scores (n=133) and for baseline covariates (n=147,) were excluded from this analysis, leaving 13,916 hypertensive participants included in the study sample (herein called ASPREE-HTN; **Supplementary Figure S1**).

### Participant baseline characteristics

ASPREE-HTN participant (N=13,916) characteristics by baseline AHM use are presented in **Table 1**. The mean [SD] age was 75.3 [4.6] years, 55.8% were female, 9,843 (70.7%) were taking AHMs at baseline, and of the AHM users, 48.2% used ARBs, 32.4% used ACEIs, 15.5% used BBs, 35.3% used diuretics and 32.9% used CCBs. ARB use was lowest in Blacks and Hispanics/Latinos (27.6% and 27.3%, respectively), compared to Whites (50%), with CCB and diuretics more common in Blacks (44.6% and 56%, respectively) and ACEIs most common in Hispanic/Latinos (49.5%). Additionally, 99.4% (n=9,782) remained on at least one AHM at each annual visit throughout follow-up. Compared to baseline untreated HTN participants, AHM users were more likely to be female, Black, never to have consumed alcohol, had a lower average education level, lived alone, had a higher prevalence of diabetes and CKD, polypharmacy and statin use (**Table 1**). The majority were using only one AHM (49.5%), with 35% using 2 AHMs and 15.5% using ≥3. Importantly, the prevalence of family history of dementia and APOE ε4 carrier status was similar across all AHM groups. Of the ‘untreated group’ (n=4073), 50.5% were prescribed an AHM at some stage during follow-up (**Supplementary Table 1**).

**Table 1.**
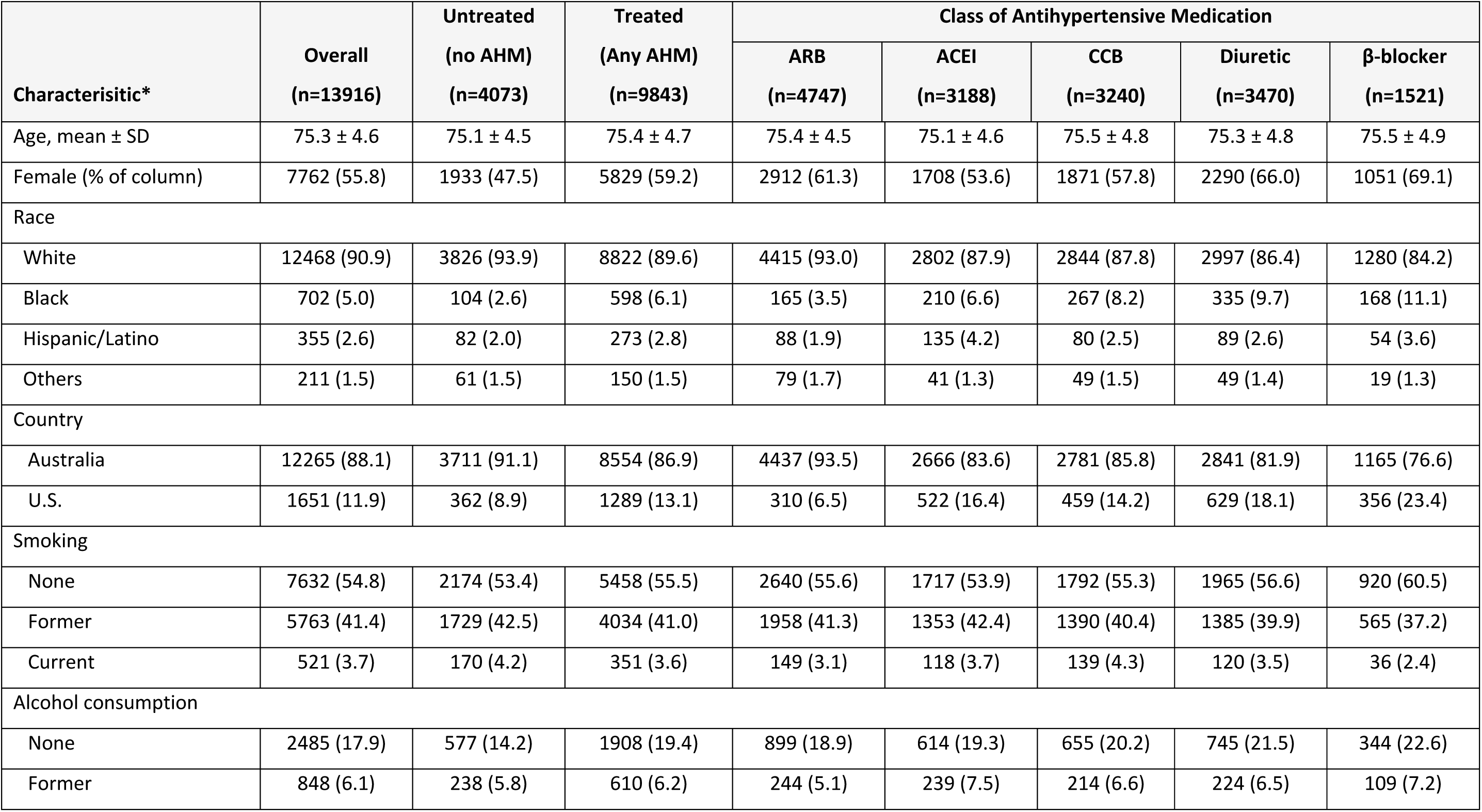
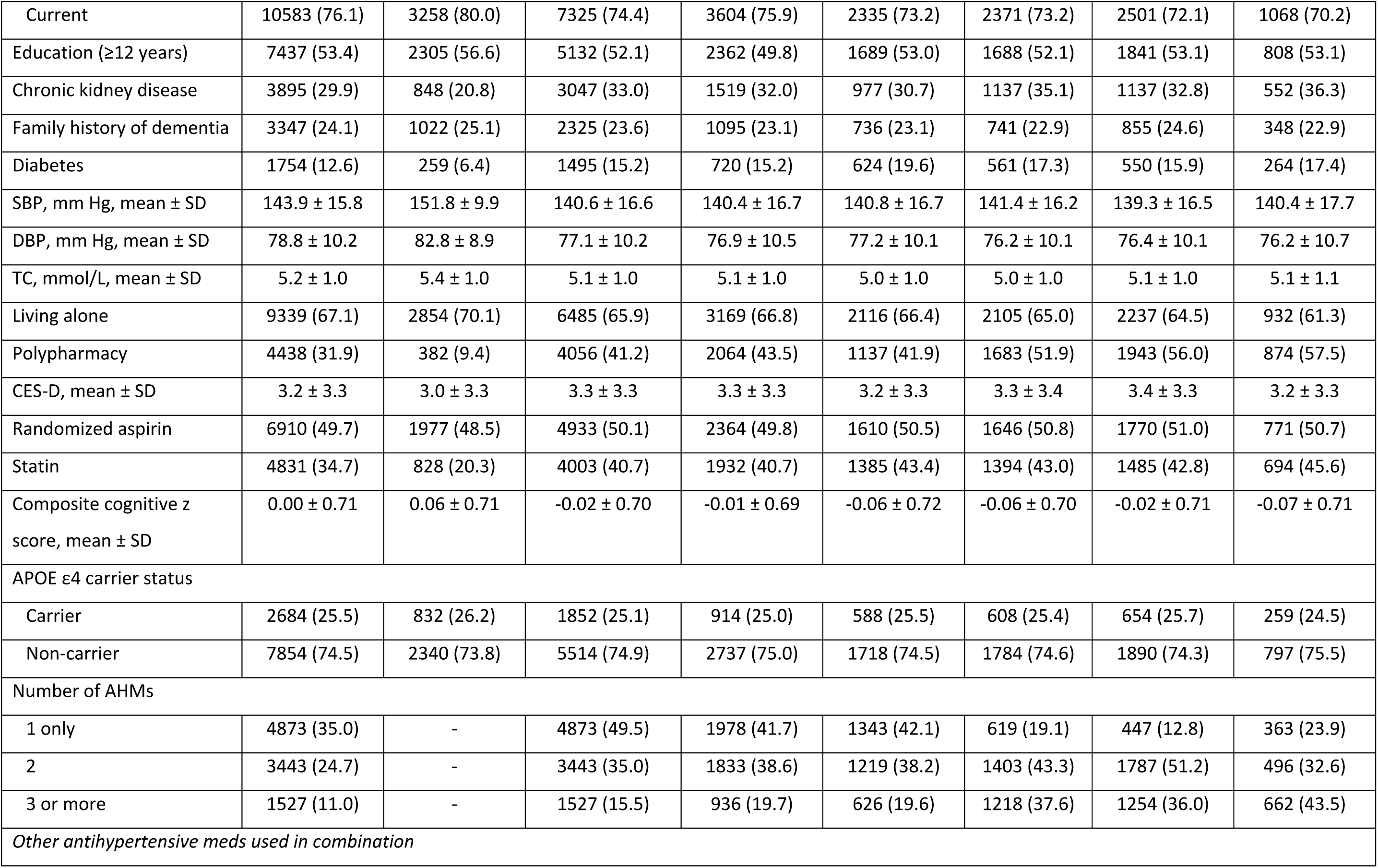
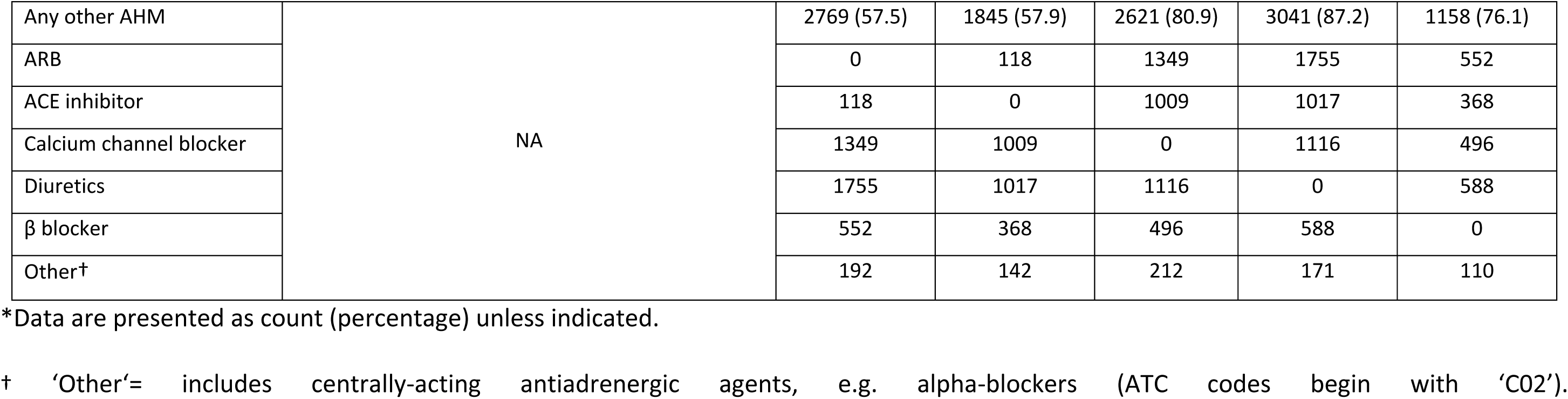
Hypertensive cohort (ASPREE-HTN) baseline characteristics between antihypertensive medication (AHM) treated (not mutually exclusive) and untreated groups.

Baseline characteristics of participants without hypertension (excluded from this analysis) or who used AHM monotherapy, are shown in **Supplementary Tables 2and 3**, respectively. Compared to those without HTN, those with HTN had higher prevalence of CKD, diabetes, statin use and polypharmacy (**Supplementary Table 2**).

### AHM use and incident dementia

Table 2 shows a comparison between specific AHM-treated hypertensive groups (as monotherapy or in combination with other AHMs) vs the untreated group, on dementia risk. During follow-up, there were 638 incident dementia cases; 193/4,073 (4.7%) in the untreated group and 445/9,843 (4.5%) in the treated group. Compared with no use, any-AHM use was not associated with a significant change in dementia risk (Model 2 [fully adjusted] HR 0.84, 95% CI 0.70-1.02; p=0.08). Adjusting for APOE ε4 carrier status or introducing a 2-year lag did not alter these findings (**Supplementary Table 4 and Table 5**).

**Table 2.**
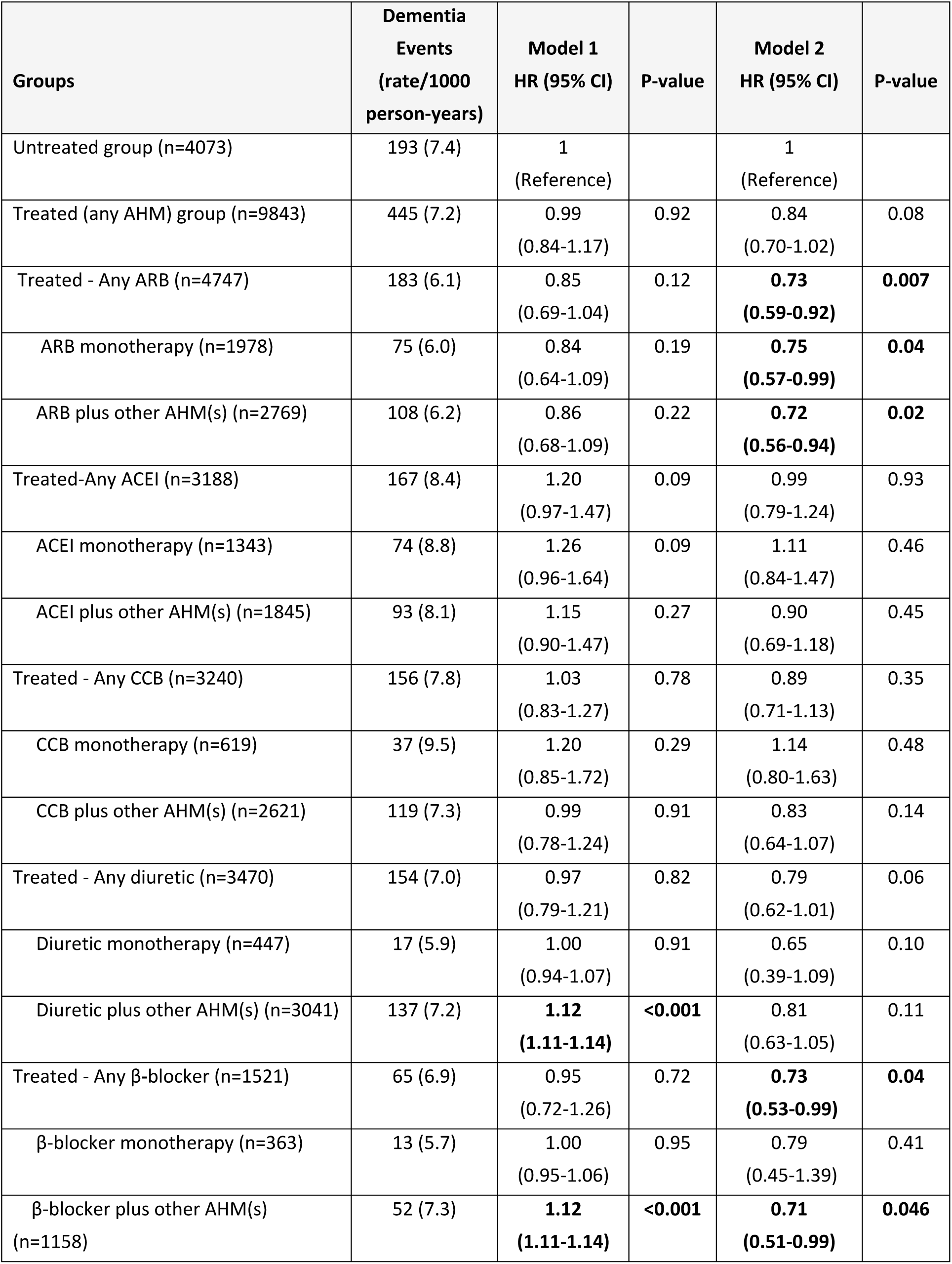
Risk of incident dementia between treated (by ‘any AHM’ and by specific AHM class) and untreated groups in participants with baseline hypertension (ASPREE-HTN population).

| Groups | Dementia Events<br>(rate/1000 person-years) | Model 1<br>HR (95% CI) | P-value | Model 2<br>HR (95% CI) | P-value |
| --- | --- | --- | --- | --- | --- |
| Untreated group (n=4073) | 193 (7.4) | 1<br>(Reference) |  | 1<br>(Reference) |  |
| Treated (any AHM) group (n=9843) | 445 (7.2) | 0.99<br>(0.84-1.17) | 0.92 | 0.84<br>(0.70-1.02) | 0.08 |
| Treated - Any ARB (n=4747) | 183 (6.1) | 0.85<br>(0.69-1.04) | 0.12 | **0.73**<br>**(0.59-0.92)** | **0.007** |
| ARB monotherapy (n=1978) | 75 (6.0) | 0.84<br>(0.64-1.09) | 0.19 | **0.75**<br>**(0.57-0.99)** | **0.04** |
| ARB plus other AHM(s) (n=2769) | 108 (6.2) | 0.86<br>(0.68-1.09) | 0.22 | **0.72**<br>**(0.56-0.94)** | **0.02** |
| Treated-Any ACEI (n=3188) | 167 (8.4) | 1.20<br>(0.97-1.47) | 0.09 | 0.99<br>(0.79-1.24) | 0.93 |
| ACEI monotherapy (n=1343) | 74 (8.8) | 1.26<br>(0.96-1.64) | 0.09 | 1.11<br>(0.84-1.47) | 0.46 |
| ACEI plus other AHM(s) (n=1845) | 93 (8.1) | 1.15<br>(0.90-1.47) | 0.27 | 0.90<br>(0.69-1.18) | 0.45 |
| Treated - Any CCB (n=3240) | 156 (7.8) | 1.03<br>(0.83-1.27) | 0.78 | 0.89<br>(0.71-1.13) | 0.35 |
| CCB monotherapy (n=619) | 37 (9.5) | 1.20<br>(0.85-1.72) | 0.29 | 1.14<br>(0.80-1.63) | 0.48 |
| CCB plus other AHM(s) (n=2621) | 119 (7.3) | 0.99<br>(0.78-1.24) | 0.91 | 0.83<br>(0.64-1.07) | 0.14 |
| Treated - Any diuretic (n=3470) | 154 (7.0) | 0.97<br>(0.79-1.21) | 0.82 | 0.79<br>(0.62-1.01) | 0.06 |
| Diuretic monotherapy (n=447) | 17 (5.9) | 1.00<br>(0.94-1.07) | 0.91 | 0.65<br>(0.39-1.09) | 0.10 |
| Diuretic plus other AHM(s) (n=3041) | 137 (7.2) | **1.12**<br>**(1.11-1.14)** | **<0.001** | 0.81<br>(0.63-1.05) | 0.11 |
| Treated - Any β-blocker (n=1521) | 65 (6.9) | 0.95<br>(0.72-1.26) | 0.72 | **0.73**<br>**(0.53-0.99)** | **0.04** |
| β-blocker monotherapy (n=363) | 13 (5.7) | 1.00<br>(0.95-1.06) | 0.95 | 0.79<br>(0.45-1.39) | 0.41 |
| β-blocker plus other AHM(s) (n=1158) | 52 (7.3) | **1.12**<br>**(1.11-1.14)** | **<0.001** | **0.71**<br>**(0.51-0.99)** | **0.046** |

When examining specific AHM classes, ARB use, as monotherapy or in combination, compared to the untreated group, was associated with a significant decrease in dementia risk; HR 0.75, 95% CI, 0.57-0.99, p=0.04 and HR 0.72, 95% CI, 0.56-0.94, p=0.02, respectively. BBs use in combination with other AHMs was significantly associated with decreased risk of dementia (‘any’ BB, HR 0.73, 95%CI, 0.53-0.99, p=0.04 and in combination with another AHM, HR 0.71, 95%CI, 0.51-0.99, p=0.046), but not for monotherapy use (HR 0.79, 95%CI, 0.45-1.39, p=0.41).

We compared specific AHM classes against each other to determine which AHM had the greatest impact on dementia risk, conducting the comparison as either ‘any’ specific AHM class (*i.e.,* monotherapy and/or combined with any other class, **Table 3A**) or as monotherapy only (**Table 3B**). ARB use, either ‘any’ or as monotherapy, was associated with a significantly lower dementia risk compared to ACEIs (by 27% and 31%, respectively) or to CCBs (by 27% and 33%, respectively). Any BB AHM use was associated with a reduction (35%) in dementia risk compared to ACEIs (**Table 3A**).

**Table 3.** Comparative risk of incident dementia in the AHM-treated hypertensive population between baseline use of different AHM classes, comparing each class (row versus column) as either ‘any’ use of the nominated class (Part A) or each class as monotherapy use (Part B).

| **A. Pairwise risk comparison between classes, with ‘any’ class use (i.e. used as monotherapy or in combination)** |  |  |  |  |  |
| --- | --- | --- | --- | --- | --- |
| **Comparison group**<br>**Treatment** | **ARB** | **ACEI** | **CCB** | **Diuretic** | **β-blocker (BB)** |
| **ARB** | 1 | 0.73<br>(0.58-0.91) | 0.73<br>(0.56-0.96) | 0.84<br>(0.62-1.15) | 0.79<br>(0.56-1.11) |
| **ACEI** | 1.37<br>(1.10-1.71) | 1 | 1.17<br>(0.90-1.52) | 1.33<br>(0.999-1.78) | 1.53<br>(1.08-2.18) |
| **CCB** | 1.37<br>(1.04-1.80) | 0.86<br>(0.66-1.11) | 1 | 1.21<br>(0.92-1.59) | 1.37<br>(0.95-1.98) |
| **Diuretic** | 1.19<br>(0.87-1.62) | 0.75<br>(0.56-1.001) | 0.83<br>(0.63-1.09) | 1 | 1.06<br>(0.73-1.52) |
| **β-blocker** | 1.26<br>(0.90-1.77) | 0.65<br>(0.46-0.93) | 0.73<br>(0.50-1.05) | 0.95<br>(0.66-1.36) | 1 |
| **B. Pairwise risk comparison between monotherapy use of different AHM classes.** |  |  |  |  |  |
| **Comparison group**<br>**Treatment** | **ARB** | **ACEI** | **CCB** | **Diuretic** | **β-blocker (BB)** |
| **ARB** | 1 | 0.69<br>(0.50-0.96) | 0.67<br>(0.45-0.996) | 1.06<br>(0.61-1.84) | 0.94<br>(0.52-1.72) |
| **ACEI** | 1.45<br>(1.04-2.01) | 1 | 0.97<br>(0.65-1.45) | 1.54<br>(0.89-2.67) | 1.37<br>(0.75-2.49) |
| **CCB** | 1.50<br>(1.003-2.24) | 1.03<br>(0.69-1.55) | 1 | 1.59<br>(0.88-2.87) | 1.41<br>(0.74-2.68) |
| **Diuretic** | 0.94<br>(0.54-1.63) | 0.65<br>(0.37-1.12) | 0.63<br>(0.35-1.13) | 1 | 0.86<br>(0.43-1.84) |
| **β-blocker** | 1.06<br>(0.58-1.93) | 0.73<br>(0.40-1.33) | 0.71<br>(0.37-1.34) | 1.13<br>(0.54-2.35) | 1 |

Participants who were taking both or none of the AHM classes involved in the comparison were excluded.
Green (HR< 1 & p<0.05) means a statistically significant lower risk of dementia in intervention group than control group, and orange (HR> 1 & p<0.05) means a statistically significant higher risk of dementia in intervention group than control group. Yellow (p≥0.05) means no statistically significant association between the two groups.

### AHM mode of action and dementia

We further explored whether AHM classes, categorized (without cross over) by their mode of action, had differential effects on dementia risk (**Table 4**). AHMs were categorized as either exclusively AT2/AT4 receptor-stimulating or -inhibiting, or by the ability to cross the blood-brain-barrier (BBB) (**Supplementary Table 6**). There was a trend towards a lower dementia risk with AT2/AT4 receptor-stimulating AHMs, compared to AT2/AT4 receptor-inhibiting AHMs, although this association was not statistically significant (HR 0.78, 95%CI 0.59-1.03, p=0.08). Within the AT2/AT4 receptor-stimulating group, ARB use was associated with a significantly decreased dementia risk compared to the AT2/AT4 receptor-inhibiting AHMs (HR 0.74, 95%CI 0.57-0.95, p=0.02), and a numerically, but not statistically significant lowered risk of dementia with thiazides (HR 0.77, 95%CI 0.56-1.05, p=0.10). BBB penetrance did not modify the association with dementia risk when comparing ARBs to ACEI classes (**Table 4**).

**Table 4.**
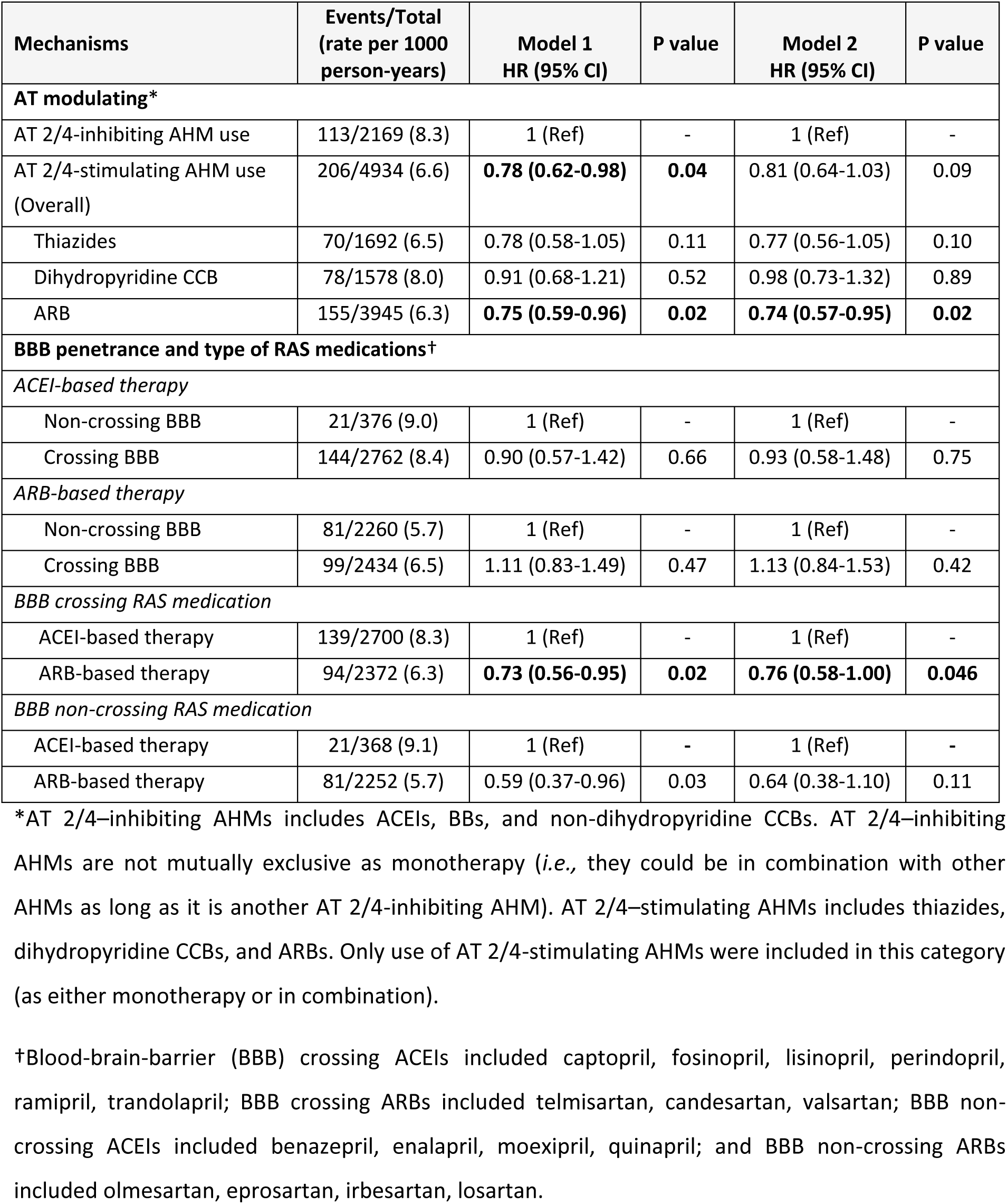
Risk of dementia between baseline use of AHMs that inhibit vs stimulate type 2 and 4 Angiotensin II receptors, or cross vs non-cross the blood-brain-barrier.

### Time-varying analysis of AHM use and incident dementia

When treating AHM class as a time-varying variable, use of ARBs, compared to other AHMs, was associated with a significant decreased dementia risk (by 24%, 31%, and 20%, for ‘any’, mono- or combo-therapy, respectively) (**Figure 1**). Conversely, ACEIs were associated with increased dementia risk by 21% for ‘any’ ACEIs, and whilst the trend for increased risk remained with mono- and combo-ACEI therapy use (by 29% and 17%, respectively), significance was lost. No other AHM class was associated with dementia risk over time.

**Figure 1.**
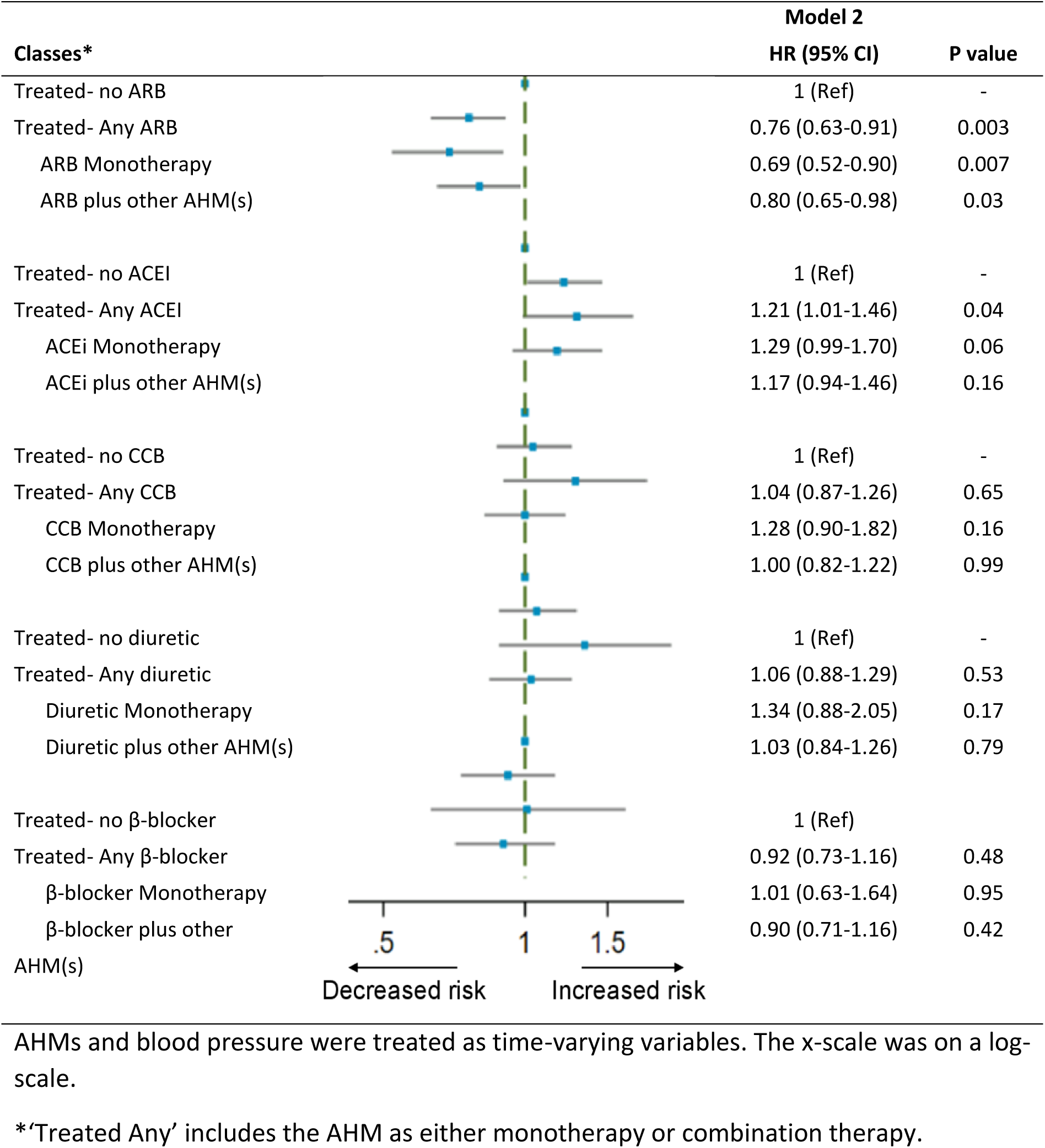
Risk of incident dementia between specific AHM treated groups (time-varying) over median 6.4 years of follow-up.

## DISCUSSION

In this post-hoc study of 13,916 hypertensive ASPREE participants followed for a median of 6.4 years, we found that compared with untreated hypertensive participants, ‘any’ baseline AHM use was not significantly associated with a change in incident dementia risk; however, when examined by AHM class, ARB use at baseline, as either combination or monotherapy, was associated with significantly decreased risk of dementia. β-Blockers were also associated with a decreased dementia risk compared to other classes, although this was only apparent when in combination with other AHMs. ARBs were associated with the greatest reduction in dementia risk compared to any other class. No other AHM class was associated with a significant change in dementia risk, although point estimates for ACEIs and CCBs trended towards harm when in monotherapy, and diuretics trended towards benefit. The time varying analysis exploring AHM exposure throughout follow-up supported the baseline exposure data, confirming the reduced dementia risk from ARB exposure at any stage during follow-up, and the increased risk with ACEIs, regardless of whether in combination or monotherapy. The differential impacts of the different AHMs account for the null finding with ‘any’ AHM use observed in this study.

Prior reports describing neuroprotection and AHMs have been inconsistent. Some observational studies and RCTs reported no significant neuroprotection with AHM use either in individuals with^31^ or without mild cognitive impairment (MCI)^32^ at baseline. Other studies have reported overall benefits of ‘any’ AHM use or class specific benefits.^33–36^ Finally, other studies have reported detrimental effects on cognition.^16,17^ Study sample size and risk profile (*e.g.,* differences in prior CVD or stroke, pre-existing hypertension, diabetes, mild cognitive decline, MCI, or dementia at baseline), comparator groups (*e.g.,* normotensive, untreated or treated hypertensive individuals, by specific AHM only), length of follow-up and differences in outcomes (MCI vs dementia) and their mode of ascertainment, may all contribute to these varied findings.^19,20^ For example, previous randomized trials reported no significant neuroprotective effects of AHM use when compared with placebo or benefits from specific AHM class in between-class comparisons.^17,37^

Whilst the strength of the evidence in these RCTs is limited by short follow-up, small sample size and younger study populations, larger more recent meta-analyses utilizing individual participant data also did not find significant AHM class differences when compared to all other AHMs combined.^21,22^ In contrast, in a large study of over 128,683 hypertensive patients with MCI, all five major classes of AHM showed a protective effect against progression to Alzheimer’s disease-related dementia (ADRD) compared to AHM users except of the AHM class being examined.^31^

Many studies have reported both ARBs and ACEIs confer similar neurocognitive benefits,^38–40^ whilst others have reported increased dementia risk with ACEIs use^41,42^ or no effect.^43,44^ Additionally, a number of large meta-analyses and studies have reported that ARBs are superior to other classes in decreasing dementia risk: when compared to diuretics, BB or CCBs^45^; when compared to all-AHMs and ACESIs^44^; and when compared to ACEIs^46^, although this last study reported BBs, CCBs and diuretics were superior to ARBs.

Our findings are in line with studies reporting neurocognitive benefits with ARBs and not ACEIs.^13,18,44,47^ We observed a 25-30% decreased dementia risk with ARB use when compared to the untreated group or directly with ACEIs (monotherapy or combination ACEIs), similar to the 20-22% risk reduction reported by others.^44,47^ When comparing ACEIs to the untreated group, we did not observe any significant outcomes, although there was a significant higher dementia risk when ACEI use was treated as a time-varying variable (P=0.04). Additionally, when ACEIs were compared to other AHM classes, a significant increased risk was observed, particularly compared to ARBs (by 37% and 45% for combination and monotherapy ACEI use, respectively).

### Mechanism

Evidence suggests that differences in dementia risk across various AHM classes may be explained by their differential impact on modulating type 2 and type 4 angiotensin II receptors and ability to penetrate the blood-brain barrier.^19,20,48^ ARBs block angiotensin-II AT1 receptors and cause upregulation of AT2 and AT4 receptors,^49^ which may promote cerebral perfusion and neurite growth, decrease vascular dysfunction and inflammation, and reduce amyloid-β and associated cholinergic deficiency–factors involved in dementia pathogenesis.^12,27^ In contrast, increased bradykinin levels resulting from ACEIs is proposed to worsen cognition, and has been linked to AD pathology.^50^ Bradykinin increases vascular permeability, stimulates prostaglandin synthesis (promoting inflammation), and increases ROS levels (associated with neuronal damage and accumulation of toxic amyloid-β).^50,51^ Higher amyloid-β accumulation has been reported in the cerebral cortex of cognitively normal adults using ACEIs, compared to ARBs users,^52^ aligning with our findings of increased dementia risk with ACEIs.

A lower risk of incident cognitive impairment and dementia has been reported with AT2/AT4 receptor-stimulating AHMs when compared with AT2/AT4 receptor-inhibiting AHMs,^19,20,46^ and with BBB-crossing RAS medications,^42^ while one study reported an increased risk with BBB-penetrance.^53^

We found AT2/AT4 receptor-stimulating AHMs compared to AT2/AT4 receptor-inhibiting AHMs, did not reach significance in reducing dementia risk, but trended to benefit (HR=0.77, 95% CI 0.56-1.05, P=0.10), reaching significance when ARBs were isolated within the AT2/AT4 receptor-stimulating group (HR=0.73, 95%CI 0.57-0.95, P=0.02). Our data suggests that AT2/AT4 receptor-stimulating AHMs may lead to decreased dementia risk compared to AT2/AT4 receptor-inhibiting AHMs, and thus, provide a mechanism of action for the neurocognitive effects of ARBs based on the angiotensin receptor hypothesis. However, we found no association with BBB penetrance, in line with other studies.^40,43^

### Study strengths and limitations

This study has several strengths. It utilizes a well described, phenotypically rich, large ‘real-world’ cohort of older adults with HTN (median age 74), without severe cognitive impairment and/or persistently high hypertension (180/105 mmHg) at enrolment and followed annually for median 6.4 years. Self-reported prescription medication use was confirmed by physical confirmation and medical record review. Dementia was a pre-specified study outcome and adjudicated by expert panel, utilizing evidentiary documentation. To remove the impact of BP and hypertension on dementia risk, we only included hypertensive participants and adjusted for baseline SBP and DBP. We compared AHM exposure to untreated and treated groups, as well as direct class-to-class comparisons and repeated this in a subgroup with known APOE e4 carrier status.

Several limitations should also be noted. This study is observational and therefore may be biased by residual confounding, in addition to possible indication bias. Neither AHM dose nor prior duration of use were recorded in ASPREE, thus, the long-term association on dementia risk of AHMs could not be explored. Subgroup analyses may only have modest power to detect associations in selected subgroups and the significance of the interactions between AHM use and stratification variables. Since all ASPREE participants were free of dementia at trial entry and were generally healthier than the wider older population, our findings cannot be generalized to subjects with either MCI or dementia, and those with major multimorbidity.

## Conclusion

In this hypertensive community-dwelling older adult population, compared with non-use, any AHM use was not associated with change in risk of incident dementia. However, specific AHM classes were associated with change in risk, and its direction (benefit vs harm) was driven by AHM type, with ARBs and diuretics associated with lower risk, and ACEIs associated with higher risk. These findings appeared to be linked to AT2/AT4 receptor-stimulating AHMs but did not differ by blood-brain barrier permeability. The study results must be interpreted with caution due to the study’s observational nature and will require confirmation by randomized clinical trials designed to explore the effects of AHMs on dementia risk in healthy older populations, in those at higher risk of dementia or with MCI and/or CVD.

## NONSTANDARD ABBREVIATIONS AND ACRONYMS

ACEI: Angiotensin Converting Enzyme inhibitor
AHM: Anti-Hypertension Medication
APOE ε4: Apolipoprotein E, variant ε4
ARBs: Angiotensin receptor blocker
ASPREE: ASPirin in Reducing Events in the Elderly
ASPREE-HTN: ASPREE Hypertension population
ASPREE-XT: ASPREE eXTension observational study
AT1/AT2/AT4: Angiotensin II receptor type 1 / 2 / 4
ATC code: Anatomical Therapeutic Chemical code
BB: Beta-Blocker
BBB: Blood-brain-barrier
CCB: Calcium Channel Blocker
CES-D 10: Centre for Epidemiologic Studies-Depression, 10 item scale
DBP: Diastolic blood pressure
HTN: Hypertension
RAS: Renin Angiotensin System
SBP: SBP = Systolic blood pressure
TC: Total cholesterol
3MS: Modified Mini Mental Examination

## ACKNOWLEDGEMENTS

Bayer AG provided aspirin and matching placebo for ASPREE. The authors acknowledge the dedicated and skilled staff in Australia and the U.S. for the conduct of the trial. The authors are also most grateful to the ASPREE participants, who so willingly volunteered for this study, and the general practitioners and medical clinics who supported ASPREE participants, endorsing organizations, the ASPREE Investigator Group and all members of the ASPREE investigator team.

## REFERENCES

1. World Health Organization. Geneva, Switzerland: World Health Organization; 2021. Fact sheets of dementia [Internet] [cited 2022 Apr 13]. Available from: https://www.who.int/news-room/fact-sheets/detail/dementia.

2. Lucca U, Tettamanti M, Tiraboschi P, Logroscino G, Landi C, Sacco L, Garrì M, Ammesso S, Biotti A, Gargantini E, et al. Incidence of dementia in the oldest-old and its relationship with age: The Monzino 80-plus population-based study. Alzheimers Dement. 2020;16(3):472–481. doi: 10.1016/j.jalz.2019.09.083

3. Abell JG, Kivimäki M, Dugravot A, Tabak AG, Fayosse A, Shipley M, Sabia S, Singh-Manoux A. Association between systolic blood pressure and dementia in the Whitehall II cohort study: role of age, duration, and threshold used to define hypertension. Eur Heart J. 2018;1;39(33):3119-3125. doi: 10.1093/eurheartj/ehy288

4. Cheng W, Du Y, Zhang Q, Wang X, He C, He J, Jing F, Ren H, Guo M, Tian J, Xu Z. Age-related changes in the risk of high blood pressure. Front Cardiovasc Med. 2022;15;9:939103. doi: 10.3389/fcvm.2022.939103

5. Mozaffarian D, Benjamin EJ, Go AS, Arnett DK, Blaha MJ, Cushman M, de Ferranti S, Despres JP, Fullerton HJ, Howard VJ, et al., American Heart Association Statistics Committee and Stroke Statistics Subcommittee. Heart disease and stroke statistics--2015 update: a report from the American Heart Association. Circulation. 2015;131:e29–322

6. Ihle-Hansen H, Thommessen B, Fagerland MW, Øksengård AR, Wyller TB, Engedal K, Fure B. Blood pressure control to prevent decline in cognition after stroke. Vasc Health Risk Manag. 2015 Jun 9;11:311–6. doi: 10.2147/VHRM.S82839

7. McEvoy LK, Fennema-Notestine C, Eyler LT, Franz CE, Hagler DJ Jr, Lyons MJ, Panizzon MS, Rinker DA, Dale AM, Kremen WS. Hypertension-related alterations in white matter microstructure detectable in middle age. Hypertension. 2015;66(2):317–23. doi: 10.1161/HYPERTENSIONAHA.115.05336

8. Shajahan S, Peters R, Carcel C, Woodward M, Harris K, Anderson CS. Hypertension and Mild Cognitive Impairment: State-of-the-Art Review. Am J Hypertens. 2024;37(6):385–393. doi: 10.1093/ajh/hpae007

9. Canavan M, O’Donnell MJ. Hypertension and Cognitive Impairment: A Review of Mechanisms and Key Concepts. Front Neurol. 2022;13:821135. doi: 10.3389/fneur.2022.821135

10. Lyon M, Fullerton JL, Kennedy S, Work LM. Hypertension & dementia: Pathophysiology & potential utility of antihypertensives in reducing disease burden. Pharmacol Ther. 2024;253:108575. doi: 10.1016/j.pharmthera.2023.108575

11. Lee CJ, Lee JY, Han K, Kim DH, Cho H, Kim KJ, Kang ES, Cha BS, Lee YH, Park S. Blood Pressure Levels and Risks of Dementia: a Nationwide Study of 4.5 Million People. Hypertension. 2022;79(1):218–229. doi: 10.1161/HYPERTENSIONAHA.121.17283

12. Kehoe PG. The coming of age of the angiotensin hypothesis in Alzheimer’s disease: progress toward disease prevention and treatment? J Alzheimers Dis. 2018;62(3):1443–1466. doi:10.3233/JAD-171119

13. Levi Marpillat N, Macquin-Mavier I, Tropeano AI, Bachoud-Levi AC, Maison P. Antihypertensive classes, cognitive decline and incidence of dementia: a network meta-analysis. J Hypertens. 2013;31(6):1073–82. doi: 10.1097/HJH.0b013e3283603f53

14. den Brok MGHE, van Dalen JW, Abdulrahman H, Larson EB, van Middelaar T, van Gool WA, van Charante EPM, Richard E. Antihypertensive Medication Classes and the Risk of Dementia: A Systematic Review and Network Meta-Analysis. J Am Med Dir Assoc. 2021;22(7):1386–1395.e15. doi: 10.1016/j.jamda.2020.12.019

15. Hajjar I, Okafor M, McDaniel D, Obideen M, Dee E, Shokouhi M, Quyyumi AA, Levey A, Goldstein F. Effects of Candesartan vs Lisinopril on Neurocognitive Function in Older Adults With Executive Mild Cognitive Impairment: A Randomized Clinical Trial. JAMA Netw Open. 2020 Aug 3;3(8):e2012252. doi: 10.1001/jamanetworkopen.2020.12252

16. Lennon MJ, Lam BCP, Lipnicki DM, Crawford JD, Peters R, Schutte AE, Brodaty H, Thalamuthu A, Rydberg-Sterner T, et al. Use of Antihypertensives, Blood Pressure, and Estimated Risk of Dementia in Late Life: An Individual Participant Data Meta-Analysis. JAMA Netw Open. 2023;6(9):e2333353. doi: 10.1001/jamanetworkopen.2023.33353

17. Anderson C, Teo K, Gao P, Arima H, Dans A, Unger T, Commerford P, Dyal L, Schumacher H, Pogue, et al; ONTARGET and TRANSCEND Investigators. Renin-angiotensin system blockade and cognitive function in patients at high risk of cardiovascular disease: analysis of data from the ONTARGET and TRANSCEND studies. Lancet Neurol. 2011;10:43–53. doi: 10.1016/S1474-4422(10)70250-7

18. Deng Z, Jiang J, Wang J, Pan D, Zhu Y, Li H, Zhang X, Liu X, Xu Y, Li Y, Tang Y; Alzheimer’s Disease Neuroimaging Initiative†. Angiotensin Receptor Blockers Are Associated With a Lower Risk of Progression From Mild Cognitive Impairment to Dementia. Hypertension. 2022;79(10):2159–2169. doi: 10.1161/HYPERTENSIONAHA.122.19378

19. van Dalen JW, Brayne C, Crane PK, Fratiglioni L, Larson EB, Lobo A, Lobo E, Marcum ZA, Moll van Charante EP, et al. Association of systolic blood pressure with dementia risk and the role of age, u-shaped associations, and mortality. JAMA Intern Med. 2022;182:142–152

20. Marcum ZA, Cohen JB, Zhang C, Derington CG, Greene TH, Ghazi L, Herrick JS, King JB, Cheung AK, Bryan N et al; Systolic Blood Pressure Intervention Trial (SPRINT) Research Group. Association of antihypertensives that stimulate vs inhibit types 2 and 4 angiotensin II receptors with cognitive impairment. JAMA Netw Open. 2022;5:e2145319. doi: 10.1001/jamanetworkopen.2021.45319

21. Peters R, Yasar S, Anderson CS, Andrews S, Antikainen R, Arima H, Beckett N, Beer JC, Bertens AS, Booth A, et al. Investigation of antihypertensive class, dementia, and cognitive decline: A meta-analysis. Neurology. 2020;94(3):e267–e281. doi: 10.1212/WNL.0000000000008732. Epub 2019 Dec 11

22. Ding J, Davis-Plourde KL, Sedaghat S, Tully PJ, Wang W, Phillips C, Pase MP, Himali JJ, Gwen Windham B, Griswold M, et al. Antihypertensive medications and risk for incident dementia and Alzheimer’s disease: a meta-analysis of individual participant data from prospective cohort studies. Lancet Neurol. 2020;19(1):61–70. doi: 10.1016/S1474-4422(19)30393-X. Epub 2019 Nov 6.

23. McNeil JJ, Woods RL, Nelson MR, Murray AM, Reid CM, Kirpach B, Storey E, Shah RC, Wolfe RS, Tonkin AM, et al; ASPREE Investigator Group. Baseline Characteristics of Participants in the ASPREE (ASPirin in Reducing Events in the Elderly) Study. J Gerontol A Biol Sci Med Sci. 2017;72(11):1586–1593. doi: 10.1093/gerona/glw342. Erratum in: J Gerontol A Biol Sci Med Sci. 2019;74(5):748. doi: 10.1093/gerona/gly278

24. McNeil JJ, Woods RL, Nelson MR, Reid CM, Kirpach B, Wolfe R, Storey E, Shah RC, Lockery JE, Tonkin AM, et al. Effect of aspirin on disability-free survival in the healthy elderly. N. Engl. J. Med. 2018; 379, 1499–1508. doi:10.1056/nejmoa1800722

25. Ernst ME, Broder JC, Wolfe R, Woods RL, Nelson MR, Ryan J, Shah RC, Orchard SG, Chan AT, Espinoza SE, et al; ASPREE Investigator Group. Health Characteristics and Aspirin Use in Participants at the Baseline of the ASPirin in Reducing Events in the Elderly - eXTension (ASPREE-XT) Observational Study. Contemp Clin Trials. 2023;130:107231. doi: 10.1016/j.cct.2023.107231

26. Teng EL, Chui HC. The Modified Mini-Mental State (3MS) examination. J Clin Psychiatry. 1987 Aug;48(8):314–8. PMID: 3611032

27. Ho JK, Nation DA. Cognitive benefits of angiotensin IV and angiotensin-(1–7): a systematic review of experimental studies. Neurosci Biobehav Rev. 2018;92:209–225. doi: 10.1016/j.neubiorev.2018.05.005

28. American Psychiatric Association. Diagnostic and Statistical Manual of Mental Disorders: DSM-IV 4^th^ Ed. American Psychiatric Association. 1994

29. Lockery JE, Broder JC, Ryan J, Stewart AC, Woods RL, Chong TT, Cloud GC, Murray A, Rigby JD, Shah R, et al; ASPREE Investigator Group, A Cohort Study of Anticholinergic Medication Burden and Incident Dementia and Stroke in Older Adults. J Gen Intern Med. 2021;36(6):1629–1637. doi: 10.1007/s11606-020-06550-2

30. Radloff LS. (1977). The CES-D Scale: A self-report depression scale for research in the general population. App. Psycolh. Meas. 1977;1,385–401

31. Lundin SK, Hu X, Feng J, Lundin KK, Chen Y, Tao C. Associations of Antihypertensive Medication Consumption and Drug-Drug Interaction with Statin and Metformin with Reduced Alzheimer’s Disease and Related Dementias Risk among Hypertensive Patients with Mild Cognitive Impairment using High Volume Claims Data. Res Sq [Preprint]. 2023:rs.3.rs-2629005. doi: 10.21203/rs.3.rs-2629005/v1

32. Peters R, Xu Y, Fitzgerald O, Aung HL, Beckett N, Bulpitt C, Chalmers J, Forette F, Gong J, Harris K, et al; Dementia rIsk REduCTion (DIRECT) collaboration. Blood pressure lowering and prevention of dementia: an individual patient data meta-analysis. Eur Heart J. 2022;43(48):4980–4990. doi: 10.1093/eurheartj/ehac584

33. Hughes D, Judge C, Murphy R, Loughlin E, Costello M, Whiteley W, Bosch J, O’Donnell MJ, Canavan M. Association of Blood Pressure Lowering With Incident Dementia or Cognitive Impairment: A Systematic Review and Meta-analysis. JAMA. 2020 May 19;323(19):1934–1944. doi: 10.1001/jama.2020.4249

34. Kjeldsen SE, Narkiewicz K, Burnier M, Oparil S. Intensive blood pressure lowering prevents mild cognitive impairment and possible dementia and slows development of white matter lesions in brain: the SPRINT Memory and Cognition IN Decreased Hypertension (SPRINT MIND) study. Blood Press. 2018;27(5):247–248. doi:10.1080/08037051.2018.1507621

35. SPRINT MIND Investigators for the SPRINT Research Group; Williamson JD, Pajewski NM, Auchus AP, Bryan RN, Chelune G, Cheung AK, Cleveland ML, Coker LH, Crowe MG, Cushman WC, et al. Effect of Intensive vs Standard Blood Pressure Control on Probable Dementia: A Randomized Clinical Trial. JAMA. 2019;321(6):553–561. doi: 10.1001/jama.2018.21442

36. Hussain S, Singh A, Rahman SO, Habib A, Najmi AK. Calcium channel blocker use reduces incident dementia risk in elderly hypertensive patients: A meta-analysis of prospective studies. Neurosci Lett. 2018;671:120–127. doi: 10.1016/j.neulet.2018.02.027

37. Bosch J, O’Donnell M, Swaminathan B, Lonn EM, Sharma M, Dagenais G, Diaz R, Khunti K, Lewis BS, Avezum A, et al; HOPE-3 Investigators. Effects of blood pressure and lipid lowering on cognition: Results from the HOPE-3 study. Neurology. 2019;92(13):e1435–e1446. doi: 10.1212/WNL.0000000000007174

38. Johnson ML, Parikh N, Kunik ME, Schulz PE, Patel JG, Chen H, Aparasu RR, Morgan RO. Antihypertensive drug use and the risk of dementia in patients with diabetes mellitus. Alzheimers Dement. 2012;8(5):437–44. doi: 10.1016/j.jalz.2011.05.2414

39. Hajjar I, Hart M, Mack W, Lipsitz LA. Aldosterone, cognitive function, and cerebral hemodynamics in hypertension and antihypertensive therapy. Am J Hypertens. 2015; 28:319–25

40. Tully PJ, Helmer C, Peters R, Tzourio C. Exploiting Drug-Apolipoprotein E Gene Interactions in Hypertension to Preserve Cognitive Function: The 3-City Cohort Study. J Am Med Dir Assoc. 2019;20(2):188–194.e4. doi: 10.1016/j.jamda.2018.08.002

41. Sink KM, Leng X, Williamson J, Kritchevsky SB, Yaffe K, Kuller L, Yasar S, Atkinson H, Robbins M, Psaty B, Goff DC Jr. Angiotensin-converting enzyme inhibitors and cognitive decline in older adults with hypertension: results from the Cardiovascular Health Study. Arch Intern Med. 2009;169(13):1195–202. doi: 10.1001/archinternmed.2009.175

42. Gao Y, O’Caoimh R, Healy L, Kerins DM, Eustace J, Guyatt G, Sammon D, Molloy DW. Effects of centrally acting ACE inhibitors on the rate of cognitive decline in dementia. BMJ Open. 2013;3(7):e002881. doi: 10.1136/bmjopen-2013-002881

43. Fazal K, Perera G, Khondoker M, Howard R, Stewart R. Associations of centrally acting ACE inhibitors with cognitive decline and survival in Alzheimer’s disease. BJPsych Open. 2017;3(4):158–164. doi: 10.1192/bjpo.bp.116.004184

44. Scotti L, Bassi L, Soranna D, Verde F, Silani V, Torsello A, Parati G, Zambon A. Association between renin-angiotensin-aldosterone system inhibitors and risk of dementia: A meta-analysis. Pharmacol Res. 2021 Apr;166:105515. doi: 10.1016/j.phrs.2021.105515

45. Stuhec M, Keuschler J, Serra-Mestres J, Isetta M. Effects of different antihypertensive medication groups on cognitive function in older patients: A systematic review. Eur Psychiatry. 2017;46:1–15. doi:10.1016/j.eurpsy.2017.07.015

46. Schroevers JL, Hoevenaar-Blom MP, Busschers WB, Hollander M, Van Gool WA, Richard E, Van Dalen JW, Moll van Charante EP. Antihypertensive medication classes and risk of incident dementia in primary care patients: a longitudinal cohort study in the Netherlands. Lancet Reg Health Eur. 2024;42:100927. doi: 10.1016/j.lanepe.2024.100927

47. Li NC, Lee A, Whitmer RA, Kivipelto M, Lawler E, Kazis LE, Wolozin B. Use of angiotensin receptor blockers and risk of dementia in a predominantly male population: prospective cohort analysis. BMJ. 2010 Jan 12;340:b5465. doi: 10.1136/bmj.b5465

48. Zhou Z, Orchard SG, Nelson MR, Fravel MA, Ernst ME. Angiotensin Receptor Blockers and Cognition: a Scoping Review. Curr Hypertens Rep. 2024;1:1–19. doi: 10.1007/s11906-023-01266-0

49. Forrester SJ, Booz GW, Sigmund CD, Coffman TM, Kawai T, Rizzo V, Scalia R, Eguchi S. Angiotensin II Signal Transduction: An Update on Mechanisms of Physiology and Pathophysiology. Physiol Rev. 2018;98(3):1627–1738. doi: 10.1152/physrev.00038.2017

50. Singh PK, Chen ZL, Ghosh D, Strickland S, Norris EH. Increased plasma bradykinin level is associated with cognitive impairment in Alzheimer’s patients. Neurobiol Dis. 2020;139:104833. doi: 10.1016/j.nbd.2020.104833

51. Ionescu-Tucker A, Cotman CW. Emerging roles of oxidative stress in brain aging and Alzheimer’s disease. Neurobiol Aging. 2021;107:86–95. doi: 10.1016/j.neurobiolaging.2021.07.014

52. Ouk M, Wu CY, Rabin JS, Edwards JD, Ramirez J, Masellis M, Swartz RH, Herrmann N, Lanctôt KL, Black SE, Swardfager W; Alzheimer’s Disease Neuroimaging Initiative. Associations between brain amyloid accumulation and the use of angiotensin-converting enzyme inhibitors versus angiotensin receptor blockers. Neurobiol Aging. 2021;100:22–31. doi: 10.1016/j.neurobiolaging.2020.12.011

53. Nagy A, Májer R, Csikai E, Dobos A, Süvegh G, Csiba L. The Correlation between Two Angiotensin-Converting Enzyme Inhibitor’s Concentrations and Cognition. Int J Environ Res Public Health. 2022;19(21):14375. doi: 10.3390/ijerph192114375

